# Lactate dehydrogenase, a Risk Factor of Severe COVID-19 Patients: A Retrospective and Observational Study

**DOI:** 10.1101/2020.03.24.20040162

**Authors:** Yi Han, Haidong Zhang, Sucheng Mu, Wei Wei, Chaoyuan Jin, Yuan Xue, Chaoyang Tong, Yunfei Zha, Zhenju Song, Guorong Gu

**Affiliations:** Emergency Department, Zhongshan Hospital, Fudan University; Department of Radiology, Renmin Hospital of Wuhan University

## Abstract

**BACKGROUND:** The World Health Organization (WHO) has recently declared coronavirus disease 2019 (COVID-19) a public health emergency of global concern. Updated analysis of cases might help identify the characteristic and risk factors of the illness severity.

**METHODS:** We extracted data regarding 47 patients with confirmed COVID-19 from Renmin Hospital of Wuhan University between February 1 and February 18, 2020. The degree of severity of COVID-19 patients (severe vs. non-severe) was defined at the time of admission according to American Thoracic Society (ATS) guidelines for community-acquired pneumonia (CAP).

**RESULTS:** The median age was 64.91 years, 26 cases (55.31%) were male of which, and 70.83% were severe cases. Severe patients had higher APACHE II (9.92 vs 4.74) and SOFA (3.0 vs 1.0) scores on admission, as well as the higher PSI (86.13 vs 61.39), Curb-65 (1.14 vs 0.48) and CT semiquantitative scores (5.0 vs 2.0) when compared with non-severe patients. Among all univariable parameters, APACHE II, SOFA, lymphocytes, CRP, LDH, AST, cTnI, BNP, et al were significantly independent risk factors of COVID-19 severity. Among which, LDH was most positively related both with APACHE II (R = 0.682) and SOFA (R = 0.790) scores, as well as PSI (R = 0.465) and CT (R = 0.837) scores. To assess the diagnostic value of these selected parameters, LDH (0.9727) had maximum sensitivity (100.00%) and specificity (86.67%), with the cutoff value of 283. As a protective factor, lymphocyte counts less than 1.045 ⨯ 10^9^ /L showed a good accuracy for identification of severe patients with AUC = 0.9845 (95%CI 0.959-1.01), the maximum specificity (91.30%) and sensitivity (95.24%). In addition, LDH was positively correlated with CRP, AST, BNP and cTnI, while negatively correlated with lymphocyte cells and its subsets, including CD3^+^, CD4^+^ and CD8^+^ T cells (P < 0.01).

**CONCLUSIONS:** This study showed that LDH could be identified as a powerful predictive factor for early recognition of lung injury and severe COVID-19 cases. And importantly, lymphocyte counts, especially CD3^+^, CD4^+^, and CD8^+^ T cells in the peripheral blood of COVID-19 patients, which was relevant with serum LDH, were also dynamically correlated with the severity of the disease.

**FUNDING:** Key Project of Shanghai Municipal Health Bureau (2016ZB0202)

## Introduction

First reported in Wuhan, Hubei province, China, on December 2019, outbreak of a viral pneumonia has attracted extensive attention of international community (1). The pathogen, a novel β-coronavirus, has currently been named severe acute respiratory syndrome coronavirus 2 (SARS-CoV-2) by the International Committee on Taxonomy of Viruses(2). The World Health Organization (WHO) has recently declared coronavirus disease 2019 (COVID-19) a public health emergency of global concern(3). As of February 29, 2020, more than 80,000 confirmed cases have been documented, with more than 10,000 severe cases, and a mortality rate of 4-15%(2, 4).

Among recent studies, the presence of any coexisting illness was more common among patients with severe disease(2). Most of the patients had elevated levels of C-reactive protein, and lymphocytopenia was common, especially in severe cases(1, 2, 5), which was thought to be a result of reduction of CD3^+^, CD4^+^ and CD8^+^ T cells(6). Meanwhile, prolonged prothrombin time (PT) and elevated lactate dehydrogenase (LDH) was found in more than 40% cases during the whole disease period(5, 7), with elevated ALT and AST less common in COVID-19 patients(2).

Given the rapid spread of COVID-19, we considered that an updated analysis of risk factors may help early recognition of the severity of the disease. In this study, we analyzed the clinical and laboratory parameters of severe and non-severe COVID-19 patients in order to evaluate disease severity.

## Results

### 1. Demographics and characteristics of COVID-19 patients

Diagnosis of COVID-19 was made according to World Health Organization interim guidance(8). A total of 47 diagnosed cases were enrolled in this study, with 24 severe cases and 23 non-severe cases (Table 1). The median age was 64.91 years (IQR, 31-87 years), 26 cases (55.31%) were male of which 70.83% were severe cases (*P* < 0.05). A total of 32 (68.09%) patients had underlying conditions, including hypertension (18 [38.3%]), diabetes (7 [14.89%]), coronary heart disease (5 [10.64%]), and autoimmune disease (2 [4.26%]); 7 (14.89%) had a history of smoking, and 6 cases (12.76%) had a BMI ≥ 25. Although no significant difference on the underlying conditions between the two groups, the patients with coronary heart disease (4, [16.67%]) and autoimmune disease (2, 8.33%) in severe group were more than the ones in non-severe patient group. All patients enrolled had fever, with maximum temperatures of approximately 38.2°C of which 12 (25.53%) had fever with dyspnea. The average days between onset of symptoms to admission ranged from 3-26 days (mean 12.51 days). Severe patients had higher APACHE II (9.92 vs. 4.74, *P* < 0.001) and SOFA (3.0 vs. 1.0, *P* < 0.001) scores on admission, as well as the higher PSI (86.13 vs. 61.39, *P* = 0.003), Curb-65 (1.14 vs. 0.48, *P* = 0.001) and CT semiquantitative scores (5.0 vs. 2.0, *P* < 0.001).

**Table 1.**
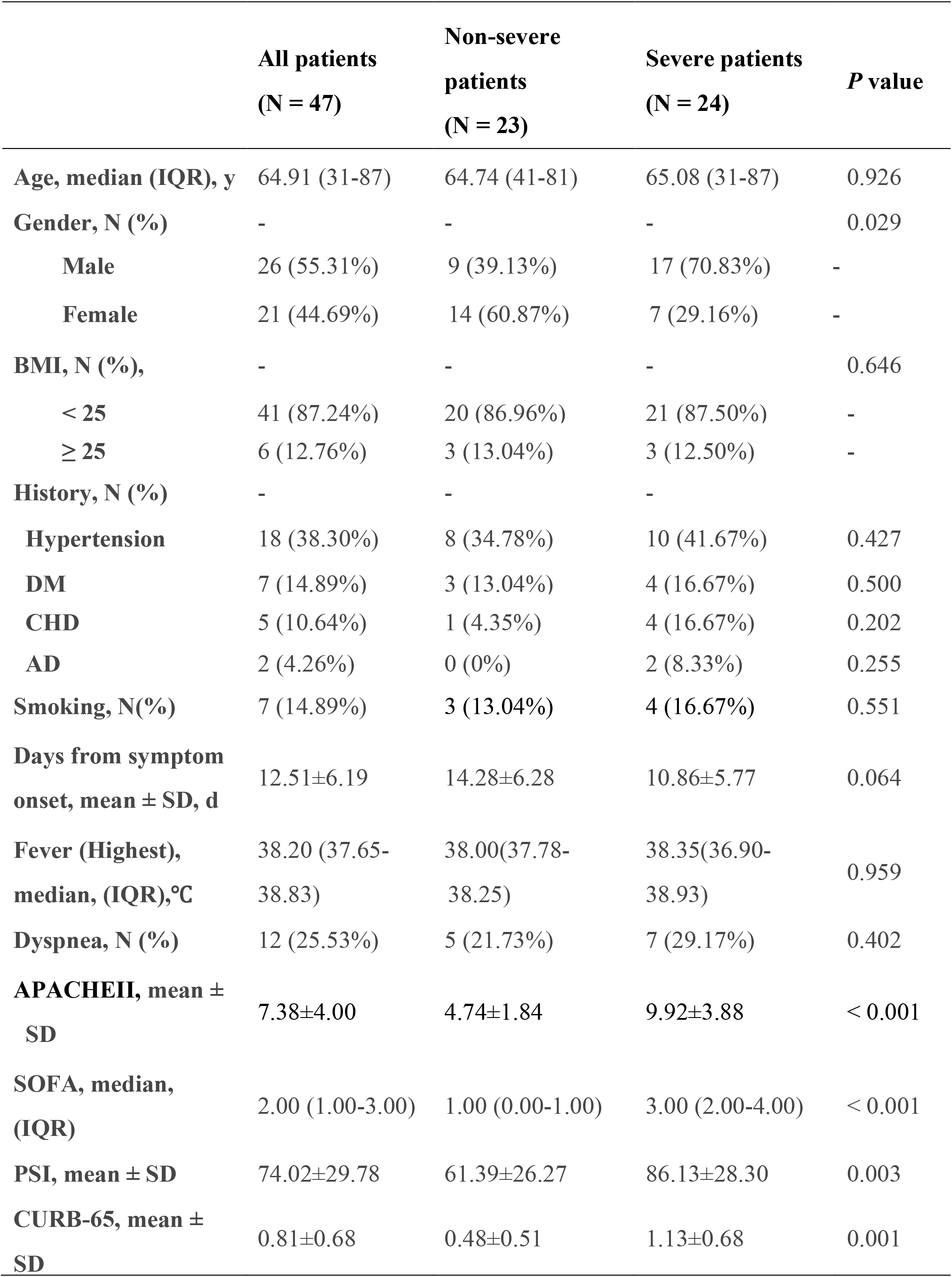
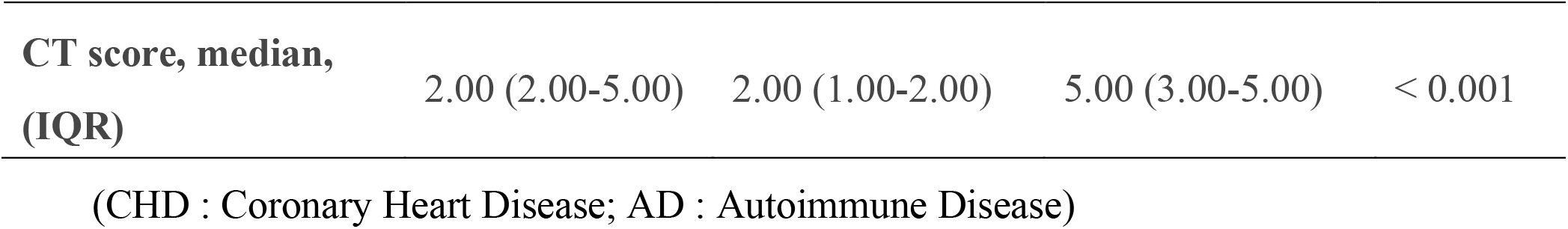
Demographic and characteristics of COVID-19 patients.

|  | **All patients**<br>**(N = 47)** | **Non-severe**<br>**patients**<br>**(N = 23)** | **Severe patients**<br>**(N = 24)** | **P value** |
| --- | --- | --- | --- | --- |
| **Age, median (IQR), y** | 64.91 (31-87) | 64.74 (41-81) | 65.08 (31-87) | 0.926 |
| **Gender, N (%)** | - | - | - | 0.029 |
| **Male** | 26 (55.31%) | 9 (39.13%) | 17 (70.83%) | - |
| **Female** | 21 (44.69%) | 14 (60.87%) | 7 (29.16%) | - |
| **BMI, N (%),** | - | - | - | 0.646 |
| **< 25** | 41 (87.24%) | 20 (86.96%) | 21 (87.50%) | - |
| **≥ 25** | 6 (12.76%) | 3 (13.04%) | 3 (12.50%) | - |
| **History, N (%)** | - | - | - |  |
| **Hypertension** | 18 (38.30%) | 8 (34.78%) | 10 (41.67%) | 0.427 |
| **DM** | 7 (14.89%) | 3 (13.04%) | 4 (16.67%) | 0.500 |
| **CHD** | 5 (10.64%) | 1 (4.35%) | 4 (16.67%) | 0.202 |
| **AD** | 2 (4.26%) | 0 (0%) | 2 (8.33%) | 0.255 |
| **Smoking, N(%)** | 7 (14.89%) | 3 (13.04%) | 4 (16.67%) | 0.551 |
| **Days from symptom**<br>**onset, mean ± SD, d** | 12.51±6.19 | 14.28±6.28 | 10.86±5.77 | 0.064 |
| **Fever (Highest),**<br>**median, (IQR),°C** | 38.20 (37.65-38.83) | 38.00(37.78-38.25) | 38.35(36.90-38.93) | 0.959 |
| **Dyspnea, N (%)** | 12 (25.53%) | 5 (21.73%) | 7 (29.17%) | 0.402 |
| **APACHEII, mean ±**<br>**SD** | 7.38±4.00 | 4.74±1.84 | 9.92±3.88 | < 0.001 |
| **SOFA, median,**<br>**(IQR)** | 2.00 (1.00-3.00) | 1.00 (0.00-1.00) | 3.00 (2.00-4.00) | < 0.001 |
| **PSI, mean ± SD** | 74.02±29.78 | 61.39±26.27 | 86.13±28.30 | 0.003 |
| **CURB-65, mean ±**<br>**SD** | 0.81±0.68 | 0.48±0.51 | 1.13±0.68 | 0.001 |
| **CT score, median,**<br>**(IQR)** | 2.00 (2.00-5.00) | 2.00 (1.00-2.00) | 5.00 (3.00-5.00) | < 0.001 |

(CHD : Coronary Heart Disease; AD : Autoimmune Disease)

### 2. Laboratory indices of COVID-19 patients

Compared to the non-severe patients, neutrophil levels (*P* < 0.001), alanine transaminase (ALT) (*P* = 0.001), aspartate transaminase (AST) (*P* < 0.001), LDH (*P* < 0.001), urea (*P* = 0.004), C-reactive protein (CRP) (*P* < 0.001), troponin I (cTnI) (P = 0.002), creatine kinase-MB (CKMB) (*P* = 0.002), B-type natriuretic peptide (BNP) (*P* = 0.006), activated partial thromboplastin time (APTT) (*P* = 0.004), D-dimer (*P* = 0.003) in severe patients were significantly higher at admission. And lymphocyte (*P* < 0.001), monocyte (*P* < 0.001), CD3^+^ (*P* < 0.001), CD4^+^ (*P* < 0.001) and CD8^+^ (*P* < 0.001) T cells in severe patients were significantly lower (Table 2). No significant differences in the serum levels of immunoglobulins (IgA, IgE, IgG and IgM) or complement C3 and C4 were observed between the two groups (Table 2).

**Table 2.** Laboratory Indices of COVID-19 patients.

|  | All patients<br>(N = 47) | Non-severe<br>patients<br>(N = 23) | Severe patients<br>(N = 24) | *P* value |
| --- | --- | --- | --- | --- |
| **White blood cell (\*10<sup>9</sup>/L)** | 7.05±3.61 | 5.41±1.67 | 8.48±4.23 | 0.022 |
| **Neutrophil (\*10<sup>9</sup>/L)** | 5.28±3.74 | 3.03±1.53 | 7.34±4.10 | < 0.001 |
| **Lymphocyte (\*10<sup>9</sup>/L)** | 1.13±0.56 | 1.57±0.48 | 0.72±0.21 | < 0.001 |
| **Monocyte (\*10<sup>9</sup>/L)** | 0.49±0.24 | 0.61±0.24 | 0.39±0.20 | < 0.001 |
| **CD3 (/μL)** | 612.63±364.42 | 973.46±298.92 | 378.10±142.33 | < 0.001 |
| **CD4 (/μL)** | 280±227.75 | 610.15±178.30 | 230.50±86.68 | < 0.001 |
| **CD8 (/μL)** | 212.26±173.78 | 345.00±194.10 | 125.98±84.71 | < 0.001 |
| **CD19 (/μL)** | 141.93±82.86 | 167.03±94.32 | 124.59±62.64 | 0.584 |
| **CD16+56 (/μL)** | 140.09±82.86 | 179.85±80.27 | 115.65±76.06 | 0.027 |
| **ALT (U/L)** | 27.0 (17.25-49) | 19.0 (16.75-28.50) | 42.0 (25.50-79.50) | 0.001 |
| **AST (U/L)** | 39.45±23.53 | 24.50±13.30 | 54.41±22.09 | < 0.001 |
| **TB (umol/L)** | 15.83±10.49 | 14.81±1.32 | 16.86±12.93 | 0.523 |
| **LDH (U/L)** | 357.85±169.56 | 228.27±67.47 | 500.40±127.19 | < 0.001 |
| **UREA (mmol/L)** | 5.56±2.36 | 4.55±1.32 | 5.56±2.75 | 0.004 |
| **Crea (umol/L)** | 64.34±22.54 | 59.09±18.29 | 69.59±25.45 | 0.124 |
| **BG (mmol/L)** | 5.62 (4.98-7.85) | 5.17 (4.84-6.04) | 6.69 (5.44-9.35) | 0.011 |
| **CRP (mg/L)** | 57.52±62.42 | 17.99±25.87 | 99.14±62.89 | < 0.001 |
| **cTnI (ng/mL)** | 0.01 (0.00-0.03) | 0.00 (0.00-0.01) | 0.01 (0.00-0.09) | 0.002 |
| **CKMB (ng/mL)** | 1.35 (0.76-2.51) | 1.19 (0.57-1.37) | 1.94 (0.92-3.33) | 0.002 |
| **BNP (pg/mL)** | 321.97±413.64 | 132.83±128.82 | 501.16±506.82 | 0.006 |
| **PT (s)** | 12.29±1.06 | 11.90±0.922 | 12.69±1.07 | 0.016 |
| **APTT (s)** | 27.7 (25.88-29.93) | 26.3 (24.85-28.35) | 28.8 (27.35-32.30) | 0.004 |
| **D-Dimer**<br>**(mg/L)** | 1.07 (0.48-4.93) | 0.64 (0.46-1.16) | 2.30 (0.86-27.15) | 0.003 |
| **Fib (g/L)** | 4.69 (3.63-6.07) | 4.31 (3.07-6.03) | 4.97 (4.50-6.30) | 0.064 |
| **IgA (g/L)** | 2.69±1.00 | 1.01±0.64 | 3.08±1.08 | 0.013 |
| **IgE (IU/mL)** | 23.25 (0.00-84.13) | 0.00 (0.00-33.98) | 36.00 (0.00-103.5) | 0.091 |
| **IgM (g/L)** | 1.01±0.39 | 1.01±0.44 | 1.01±0.36 | 0.938 |
| **IgG (g/L)** | 11.3 (9.42-14.15) | 11.05 (9.34-12.53) | 12.50 (9.82-15.20) | 0.353 |
| **C3 (g/L)** | 1.05±0.22 | 1.04±0.22 | 1.06±0.22 | 0.846 |
| **C4 (g/L)** | 0.29±0.15 | 0.32±0.20 | 0.27±0.09 | 0.383 |

### 3. Independent risk factors of severe COVID-19 patients

To assess the risk factors of the demographic, characteristics and laboratory indicators on the severity of COVID-19 patients, logistic regression analysis was performed on the parameters of significant difference using *t* test. In univariable analysis, odds ratio of SOFA score, Curb-65 score, serum CKMB concentration and serum IgA level were the highest in severe patients. Male patients infected with SARS-cov-2 showed as an independent risk factor for getting severer condition as 3.76 (1.12-12.73). Apart from the risk factors above, APACHE II score, PSI score, white blood cell count, neutrophil count, serum AST, ALT, LDH, Urea, CRP, BNP level, PT, APTT were all associated with the severity of COVID-19 patients. Meanwhile, we found that the lymphocyte counts, monocyte counts, CD3^+^, CD4^+^, CD8^+^, and CD16^+^CD56^+^ lymphocyte counts were protective factors (OR < 1) for COVID-19 patients. Based on the condition we mentioned, laboratory indictors CRP, AST and LDH (*P* < 0.001) were chosen as the three variables for multivariable logistic regression model. As a result, serum LDH level was ruled out as a risk factor for the severity of COVID-19 patients (OR:1.02, 95% CI:1.01-1.03, *P* < 0.05) (Table 3).

**Table 3.**
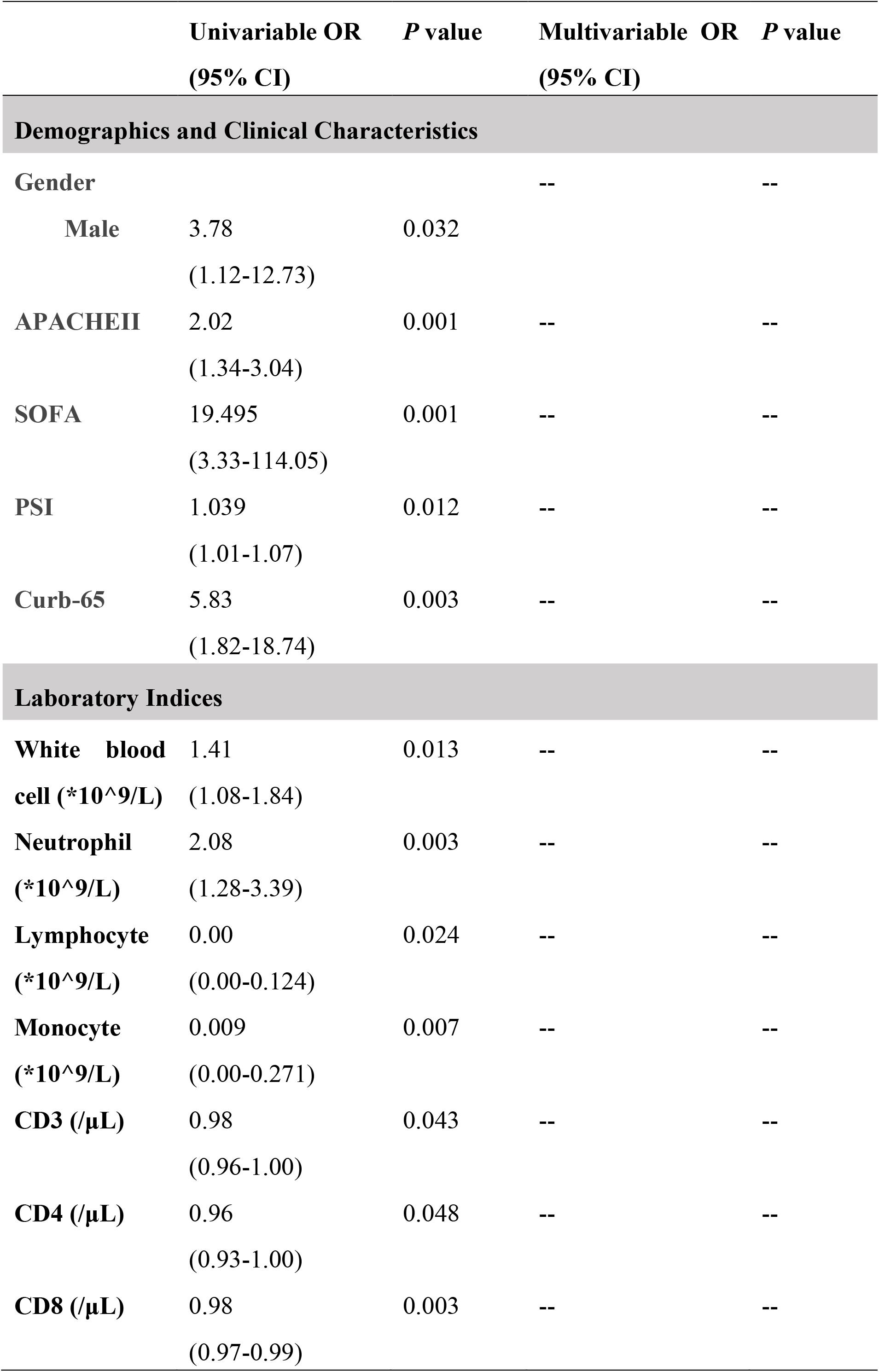
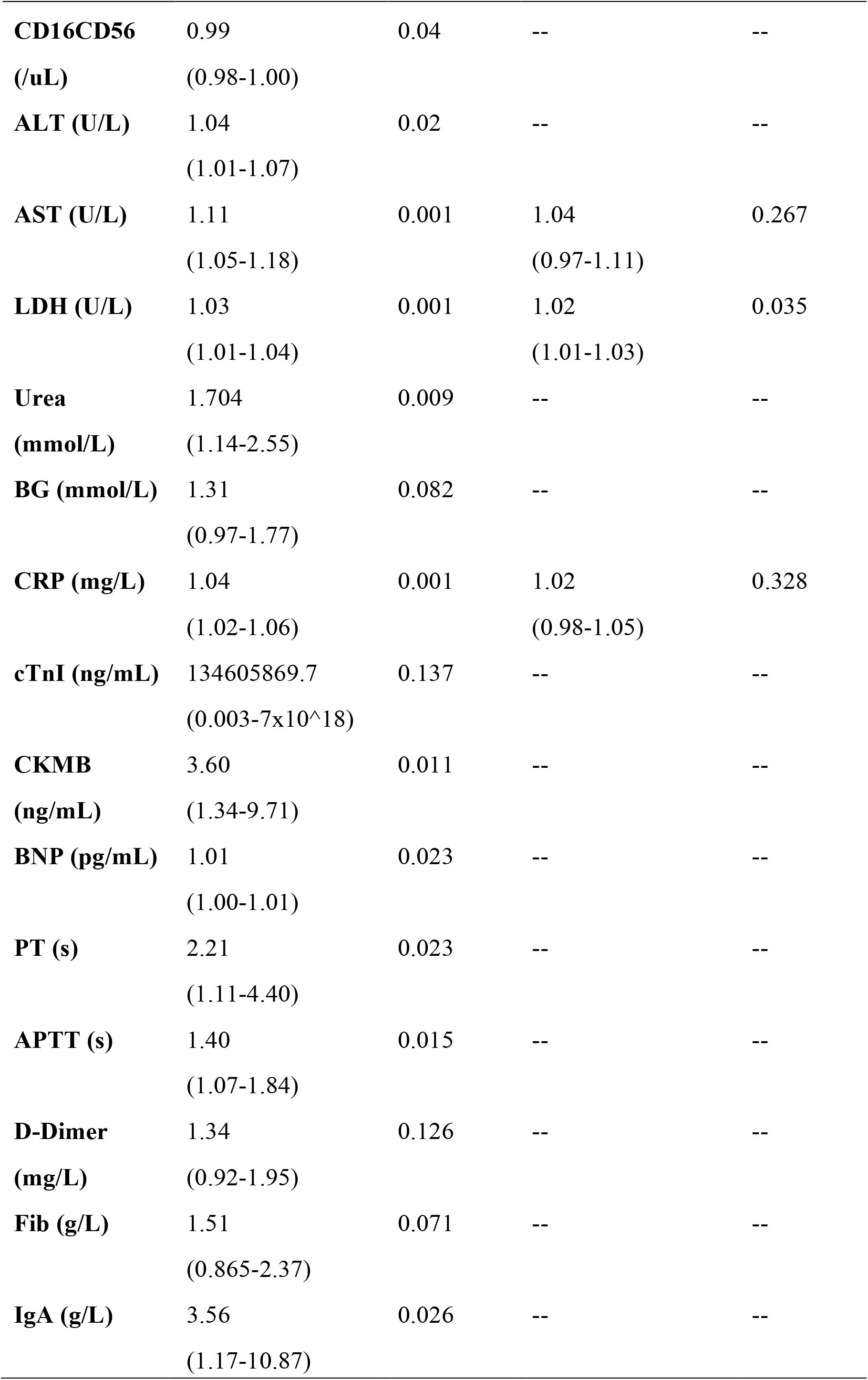
Univariable OR and Multivariable OR of severe COVID-19 patients.

### 4. The Predictive factors correlated with severity of COVID-19

We used clinical severity scores (APACHE II and SOFA) to assess the disease severity in COVID-19 patients. The average APACHE II score was 9.9 in severe cases versus 4.7 in non-severe cases, with SOFA score of 3.0 versus 1.0 (Table 1, *P* < 0.001). For the predivtive factors we have speculated, lymphocyte cell counts, hepatic function indicator AST, infection indicator CRP and myocardial injury biomarkers cTnI, BNP as well as LDH were performed Pearson correlation ananalysis with APACHE II score and SOFA score. We found that serum LDH level showed tightly positive correlation of the highest R value respevtively with APACHE II (R = 0.682, *P* < 0.001) and SOFA score (R = 0.790, *P* < 0.001) in all the indicators (Figure1. F&). Apart from lymphocyte cells showed a negative correlation with APACHE II (R = −0.593, *P* < 0.001) and SOFA score (R = −0.614, *P* < 0.001), other indicators CRP, AST and BNP were also positively associated with the scores. It’s worth noting that, the serum cTnI level showed a positive correlation with SOFA score (R = 0.345, *P* = 0.025) while it showed no correlation with APACHE II score (R = 0.292, *P* = 0.06). (Figure 1).

**Figure 1.**
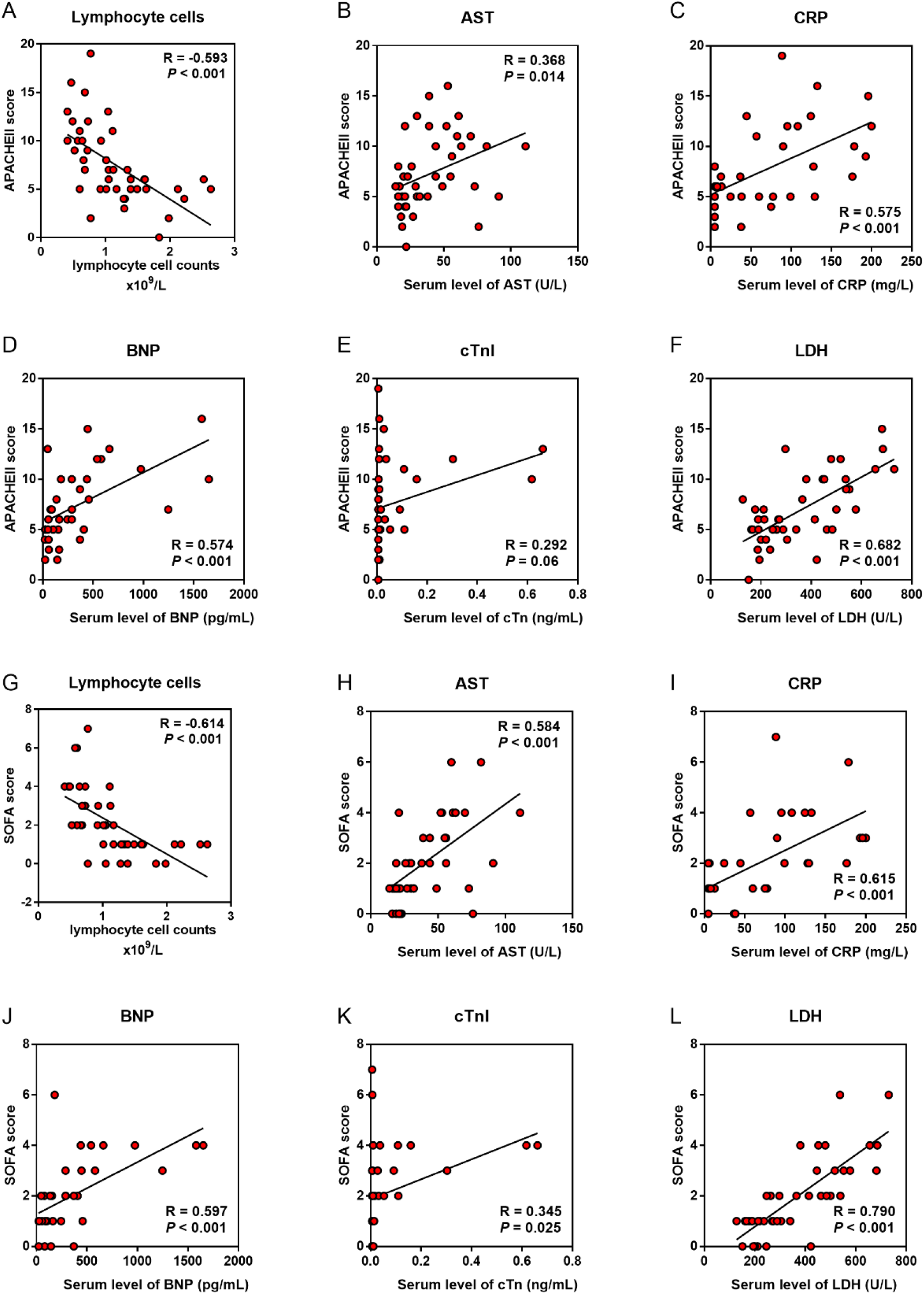
Predictive factors correlated with severity of COVID-19 patients. (A-F: Pearson correlation analysis was performed between candidate indicators with APACHEII score. G-L: Pearson correlation analysis was performed between indicators with SOFA score.)

### 5. The predictive factors correlated with the severity of lung damage

We used PSI and the Curb-65 score to assess the disease severity of pneumonia in COVID-19 patients. The average PSI score was 86 in severe cases versus 61 in non-severe cases (Table 1, *P* = 0.003), with Curb-65 scores of 1.13 versus 0.48 (Table 1, *P* = 0.001). We also evaluated the extent of inflammation on chest CT using a semiquantitative scoring system (Supplement Table 1); the median score was 5.0 in severe cases while only 2.0 in non-severe cases (Table 1, *P* < 0.001). Six indicators above were futher performed Pearson correlation ananalysis with PSI score and CT score to determine the potential biomarkers for the lung injury. As a result, the serum LDH level still showed the highest R value with CT score (R = 0.8372, *P* < 0.001) and the second highest R value with PSI score (R = 0.465, *P* < 0.001) in all the indicators (Figure2. F&). Nevertheless, the serum cTnI level showed no correlation with PSI score (R = 0.203, *P* = 0.198) nor CT score (R = 0.232, *P* = 0.167). What’s more, the serum AST level showed no correlation with PSI score (R = 0.211, *P* = 0.165) and the serum BNP level showed no correlation CT score (R = 0.301, *P* = 0.837). Lymphocyte counts were neagtively associated with PSI score (R = −0.434, *P* = 0.003) and CT score (R = −0.602, *P* < 0.001), while serum CRP showed positvie correlation with the two scores. (Figure 2)

**Figure 2.**
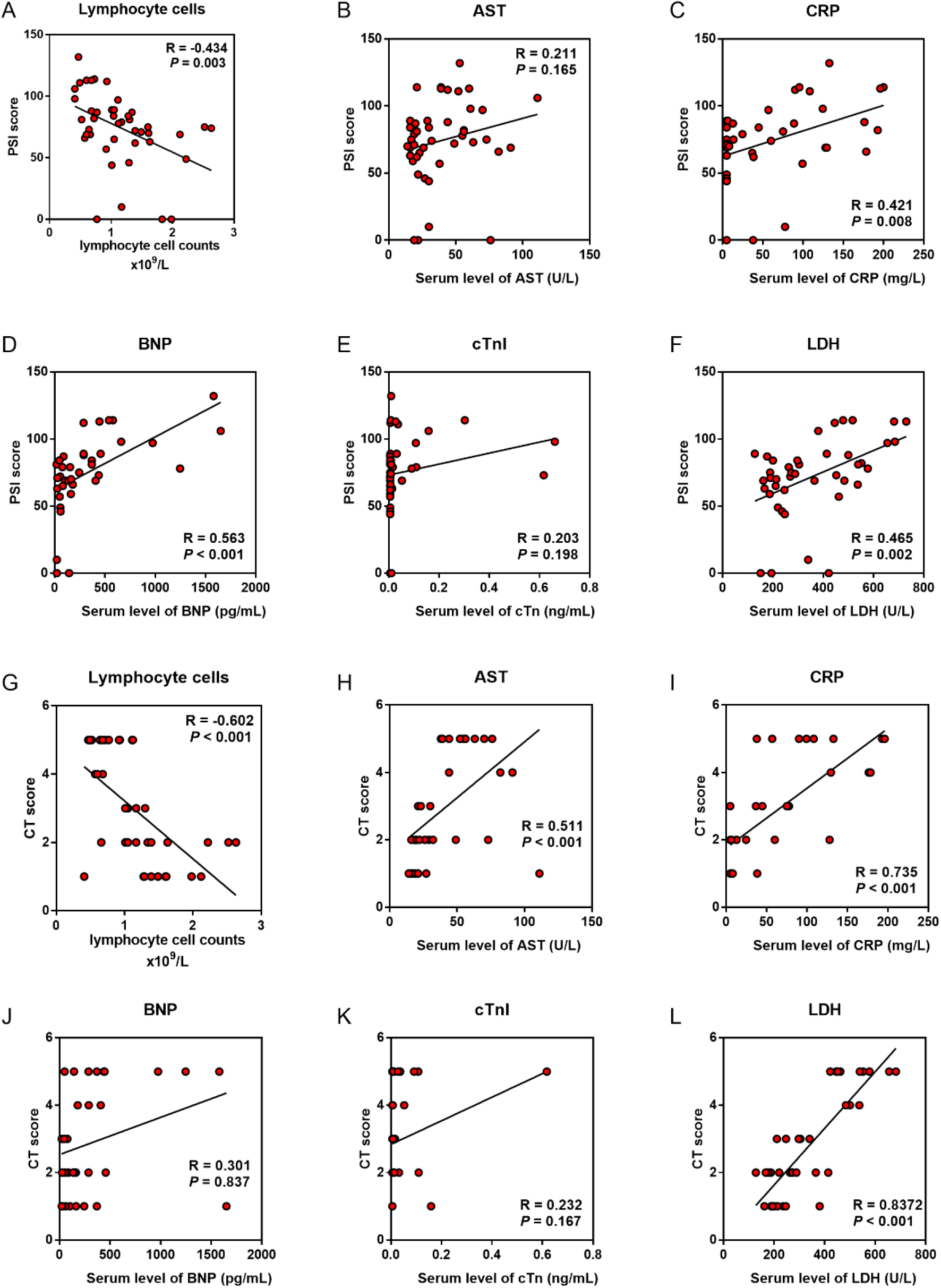
Predictive factors correlated with lung injury of COVID-19 patients. (A-F: Pearson correlation analysis was performed between the indicators with PSI score. G-L: Pearson correlation analysis was performed between the indicators with CT score.)

**Figure 3.**
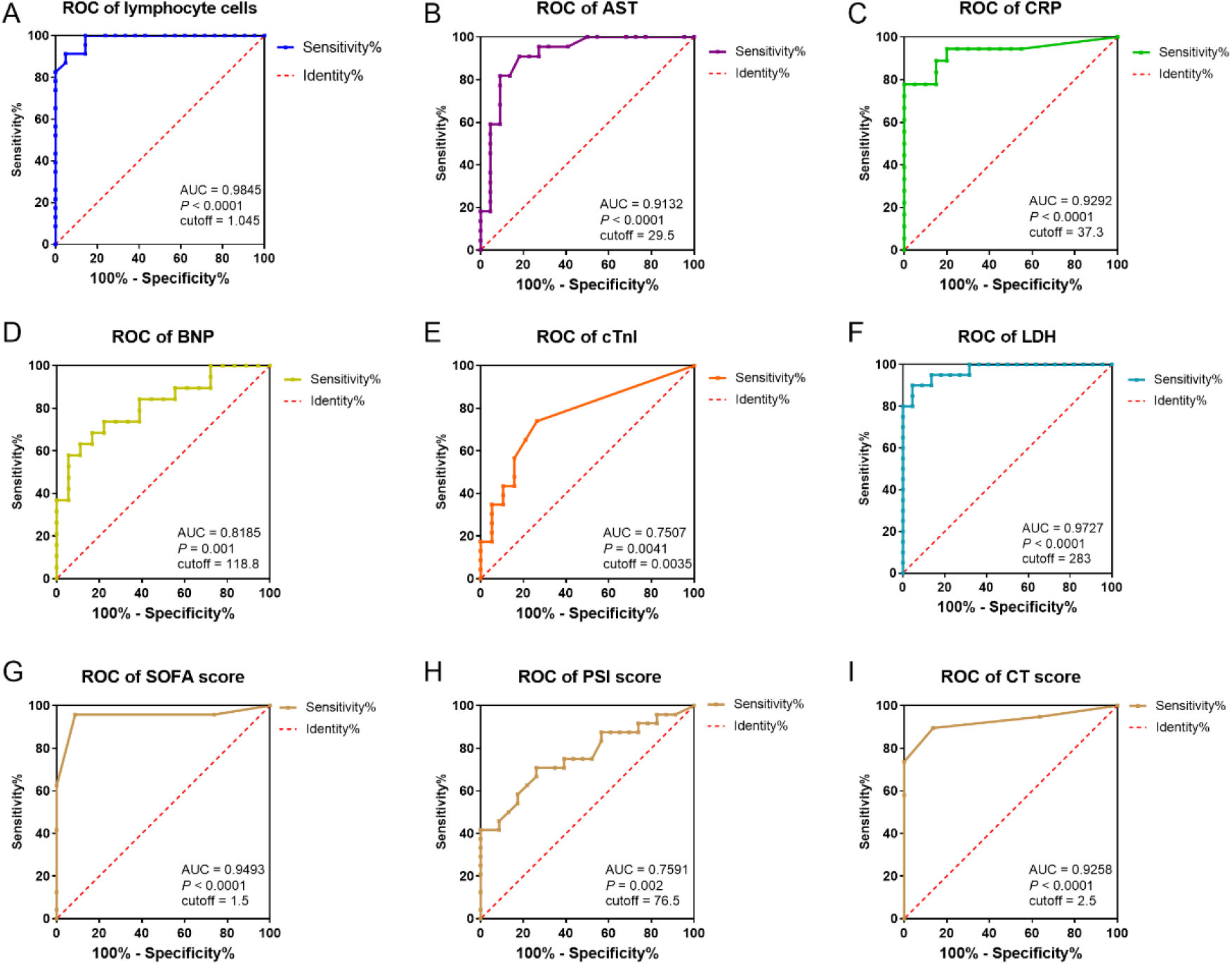
ROC curve and cutoff value of prognostic factors. (A-I: The factors for the predictor of COVID-19 patients getting severe condition. AUC, area under curve.)

### 6. The predictive factors for identification of severe COVID-19 cases

To assess the diagnostic value of these selected parameters, receiver operating characteristic (ROC) curve and area under ROC curve (AUC) were calculated using R package “pROC”. As indicated in Figure3, the area under curve (AUC = 0.9727, 95% CI: 0.932-1.013) implied a perfect accuracy of the serum LDH level more than 283 U/L in COVID-19 patients as a predictive factor for identification of severe condition, with the maximum sensitivity (100.00%) and specificity (86.67%). The serum AST level over 29.5 U/L and CRP over 37.3 mg/L showed relative moderate accuracy with AUC = 0.9231 and AUC = 0.9292. As a protective factor, the lymphocyte counts less than 1.045 × 10^9^ /L showed a good accuracy for identification of severe patients with AUC = 0.9845 (95%CI: 0.959-1.01), the maximum sensitivity (95.24%) and specificity (91.30%). Furthermore, AUC of SOFA score is 0.9493 and CT score is 0.9528. The other indicators were relatively poor accuracy factors in ROC curve analysis (Figure3. Table 4).

**Table 4.** Cutoff Value of Predictive Factors.

|  | Sensitivity | Specificity | Cutoff Value | AUC | 95% CI | P value |
| --- | --- | --- | --- | --- | --- | --- |
| Lymphocytes | 0.9524 | 0.9130 | 1.045 | 0.9845 | 0.959-1.01 | <0.0001 |
| CRP | 0.8571 | 0.9333 | 37.3 | 0.9292 | 0.837-1.02 | <0.0001 |
| AST | 0.8571 | 0.9333 | 29.5 | 0.9132 | 0.824-1.00 | <0.0001 |
| cTn | 0.7857 | 0.7333 | 0.0035 | 0.7597 | 0.612-0.908 | 0.0041 |
| BNP | 0.7857 | 0.6667 | 118.8 | 0.8158 | 0.679-0.952 | 0.001 |
| LDH | 1 | 0.8667 | 283 | 0.9727 | 0.932-1.013 | <0.0001 |
| CT score | 0.8947 | 0.8636 | 2.5 | 0.9258 | 0.8315-1.02 | <0.0001 |
| SOFA score | 0.947 | 0.909 | 1.5 | 0.9493 | 0.875-1.02 | <0.0001 |
| PSI core | 0.7083 | 0.7091 | 76.5 | 0.7591 | 0.620-0.898 | 0.002 |

### 7. Relationship between LDH and inflammation, cardiac and liver injury biomarkers

As the serum LDH level showed the highest Pearson Correlation Coefficient in the correlation with APACHE II score, SOFA score, PSI score and CT score, the best accuracy identification for predicting severe patients, we evaluated the relationship between LDH and lymphocyte cells number (including subsets), serum CRP, AST, BNP and cTnI level. We found LDH was positively correlated with CRP, AST, BNP and cTnI, while negatively correlated with lymphocyte cells and its subsets, including CD3^+^, CD4^+^ and CD8^+^ T cells (P < 0.01) (Figure 4).

**Figure 4.**
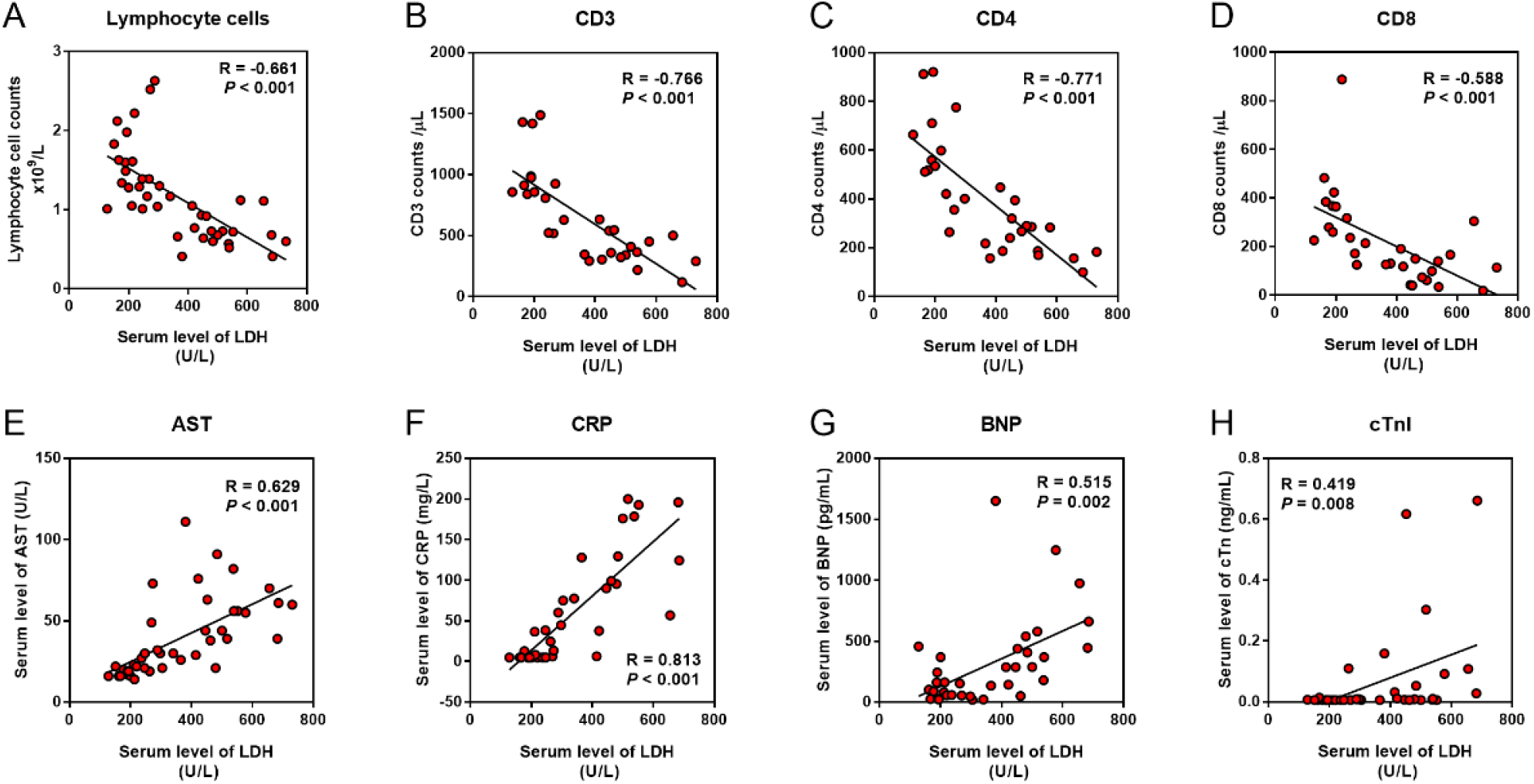
Relationship between LDH and inflammation, cardiac and liver injury. (A-H: Pearson correlation analysis was performed between the indicators with the serum LDH level.)

## Discussion

In this study, we analyzed the clinical features in 47 patients with COVID-19 who were admitted to Renmin Hospital of Wuhan University between February 1 and February 1, 2020. Although the clinical characteristics of patients enrolled were somewhat akin to those reported in previous studies(1, 5, 7), there were no differences in age, BMI, and proportion with underlying diseases between severe and non-severe patients, as well as smoking history. In concert with recent studies(1, 5, 9), we found that the clinical characteristics of COVID-19 mimic those of SARS-CoV. Fever and cough were the dominant symptoms, and both groups had similar days from symptom onset, similar maximum temperature, and number of patients who had dyspnea. APACHE II and SOFA scores were calculated based on admission data. PSI, CURB-65, and the semiquantitative score were used to assess the severity of lung damage. Significantly higher scores were found in severe cases.

Among the risk factors we investigated in this study, we surprisingly discovered that LDH had the most positive relationship between both PSI and CT score. In addition, it was also most positively relevant to APACHE II and SOFA scores, which reflected a strong correlation between LDH with lung damage as well as disease severity. LDH is a major player in glucose metabolism which is present in tissues throughout the body and catalyzes pyruvate to lactate. It is released from cells upon damage of their cytoplasmic membrane(10). Previous studies also had noted the importance of LDH as an indicator of lung diseases. In a study on Epstein-Barr virus (EBV), researchers found that EBV infected B cells had more LDH transcripts than the uninfected B cells(11). In addition, the serum levels of LDH increased in PcP patients, probably was due to lung injury(12, 13). Among patients who were infected during the 2009 influenza A (H1N1) pandemic, 77.8% whose laboratory test showed LDH > 225U/L had lung involvement, without difference between adult and children(14), which indicated that LDH elevation was associated with various pathogen including virus, and was relevant to lung injury. Furthermore, there was a case reported in 2017 that a patient with human Zika virus infection had markedly elevated LDH, which was associated with 70% mortality in further Zika infected animal study. They considered LDH as an indicator of multiorgan injury, not only affecting liver or cardiac function(15).

LDH is found in all human cells, especially in myocardial and liver cells. In our study, LDH elevation was positively associated with AST, cTnI and BNP, which verified it as an isozyme of heart and liver. However, it was somewhat surprising that cTnI was neither associated with PSI nor CT semiquantitative score, while was relevant with disease severity. Similarly, BNP that had a strong correlation with APACHE II and SOFA, was not associated with CT semiquantitative score, either. Furthermore, AST which was associated with the APACHE II and SOFA score, was not related to PSI score. This rather intriguing finding might be explained by the fact that the myocardial and liver injury caused by SARS-CoV-2 might be due to the direct damage of the virus to targeted organs, while not because of hypoxia induced by lung injury. Since the outbreak of COVID-19, structural analysis of the virus has suggested that SARS-CoV-2 might be able to bind to the angiotensin-converting enzyme 2 (ACE2) receptor in humans(16, 17). The ACE2 receptor is abundantly present in the epithelia of lung and small intestine(18), which might provide possible routes of entry for SARS-CoV-2. This epithelial expression, together with its presence in vascular endothelium(18), also provides a step in understanding the pathogenesis of ARDS, cardiac injury, liver injury and even MODS.

Furthermore, LDH was found to be positively associated with CRP and negatively with lymphocytes. An increase in CRP and decrease in lymphocytes were observed in severe cases during the 14-day observation period, which was consistent with findings of recent reports(1, 5, 6). In our study, the development of lymphopenia in severe patients was mainly related to the significantly decreased absolute counts of T cells, especially CD3^+^, CD4^+^, and CD8^+^T cells, but not to B cells or NK cells. The decrease of T cells in severe cases reached its trough within three days, and then slightly increased from the first week while still maintaining low levels and not recovering to the level of non-severe patients over two weeks (data not shown).

LDH is not only a metabolic but also an immune surveillance prognostic biomarker, its elevation is harbinger of negative outcome in immunosuppressive patients(19). LDH increases production of lactate, leads to enhancement of immune-suppressive cells, including macrophages and dendritic cells (DCs), and inhibition of cytolytic cells, such as natural killer (NK) cells and cytotoxic T-lymphocytes (CTLs)(10). LDH is often induced upon T cell activation and proliferation(20, 21). In a retrospective analysis of a CTLs antigen-4 antibody which could enhance T-cell activity and proliferation, the result showed that increase in LDH level was indicative of a poor outcome(22), that confirmed the inhibition effect of LDH on CTLs. Furthermore, CD4+ T cells produce less IFN-γ in the absence of LDH, demonstrating a critical role for LDH in promoting T cell responses(21).

It was also hypothesized that change in lactate modulated the inflammatory response in macrophages(23). Suppression of LDH has anti-inflammatory effects due to the downregulation of several inflammatory mediators including cytokines and NO(23). Also, significant correlations were found between LDH and cytokines/chemokines, therefore, LDH may be a useful biomarker to assist the clinician in the decision to hospitalize a child with bronchiolitis(24). In our study, lymphocytes, especially CD3^+^, CD4^+^, and CD8^+^ T cells were significantly decreased and relevant with LDH elevation. The decrease in T cell counts was strongly correlated with the severity of disease, which was in keeping with previous studies on SARS(25, 26). On the other hand, elevation of LDH, the immune-related factor, could be considered as a predictive factor, that reflected a poor prognosis in severe COVID-19 patients.

Our study had some limitations. This study was conducted at a single-center with limited sample size. Furthermore, because many patients remained in hospital and outcomes were unknown at the time of writing, we only collected clinical data within two weeks for our analysis. COVID-19 has spread rapidly and has a wide spectrum of severity. A larger cohort study of patients with COVID-19 pneumonia globally would help to further define the clinical characteristics and risk factors of the disease.

## Conclusion

In summary, this study showed that LDH coule be identified as a powerful predictive factor for early recognition of lung injury and severe COVID-19 cases. And importantly, lymphocyte counts, especially CD3^+^, CD4^+^, and CD8^+^ T cells in the peripheral blood of COVID-19 patients,which was relevant with serum LDH, were also dynamically correlated with the severity of the disease.

## Methods

### 1. Data collection

Forty-seven confirmed COVID-19 patients at Renmin Hospital of Wuhan University between February 1 to February 18, 2020 were enrolled into this retrospective observational study. A confirmed case of COVID-19 was defined as a positive result on real-time reverse-transcriptase-polymerase-chain-reaction (RT-PCR) assay of nasal-pharyngeal swab specimens.

A trained team of physicians and medical students reviewed and collected demographic, epidemiological, clinical, physical examination findings, and laboratory data from electronic medical records. Laboratory assessments consisted of complete blood count, liver and renal function, markers of cardiac injury, measures of electrolytes, C-reactive protein and procalcitonin, and assessment of coagulation and lactate dehydrogenase, among other parameters. We defined the degree of severity of COVID-19 patients (severe vs. non-severe) at the time of admission, according to American Thoracic Society (ATS) guidelines for CAP(27). If imaging scans were available, the radiologic assessments of chest computed tomography (CT) were reviewed and scored by an attending radiologist who extracted the data. The APACHE II and SOFA score were calculated basen on clinical and experimental data on admission, and CT score was calculated based on a semiquantitative scoring system (Supplement Table 1).

Patients were followed up for 14 days after admission. Patient information was confidentially protected by assigning a deidentified ID to each patient. The study was approved by the Ethics Committee of Renmin Hospital of Wuhan University.

### 2. Statistical Analysis

A sample size of at least 17 patients per group is needed to achieve 91% power to detect a difference of 0.3 between the area under the ROC curve (AUC) under the null hypothesis of 0.5 and an AUC under the alternative hypothesis of 0.8 using a two-sided test at a significance level of 0.05 (PASS 15, NCSS, LCC).

Because the patients enrolled in our study were not randomly assigned, all statistical findings should be interpreted as descriptive only. Quantized variables were presented as means ± standard deviation, and significance was tested by t-test. Nonparametric variables were expressed as medians and interquartile ranges or simple ranges as appropriate, and we used the Mann Whitney U or Kruskal Wallis tests to compare differences. Continuous and categorical variables were summarized as counts and percentages, and significance was detected by chi square or Fisher’s exact test. To explore the risk factors associated with severity of COVID-19, univariable and multivariate logistic regression models were used. Correlation analysis was performed by using Pearson Correlation Coefficient. The sensitivity and specificity of the risk factors for the patient diagnosis were represented and analyzed by receiver operating characteristic curve (ROC curve). All the analyses and figures were performed with SPSS software (Version 26) and Graphpad Prism (Version 7.0). *P* < 0.05 was considered statistically significant in all analyses.

## Data Availability

The data used to support the findings of this study are available from the corresponding author upon appropriate request.

## Authorship contributions

Y.H. collected, analyzed data and drafted the manuscript; H.Z. acquired CT image and evaluated lung infiltration by a semiquantitative scoring system; S.M. organized, analyzed and interpreted the data. Y.H., H.Z. and S.M. are co-first authors, the order of the authorship was based on their contributions to this study. G.G. designed the study, and took responsibility for the integrity of data and the accuracy of data analysis; Z.S. helped revise the manuscript; Y.Z. helped with technical support of radiology. G.G., Z.S. and Y.Z. are co-corresponding authors. W.W. and C.J. helped organize data and perform literature search; Y.X. provided the administrative, technical and material support; C.T. was the supervisor of this study, who provided essential theoretical guidance of this study.

## Data availability

The data used to surpport the findings of this study are avalable from the corresponding author upon approprite request.

## Acknowledgement

We are grateful to all the patients, doctors and nurses who participated in this study. We also would like to thank Dr. Quming Zhao from Children’s Hospital of Fudan University for his selfless help in practical questions and for insightful discussions, and Dr. Lingyu Xing from Zhongshan Hospital of Fudan University for her help with literature search. Their help was invaluable.

---

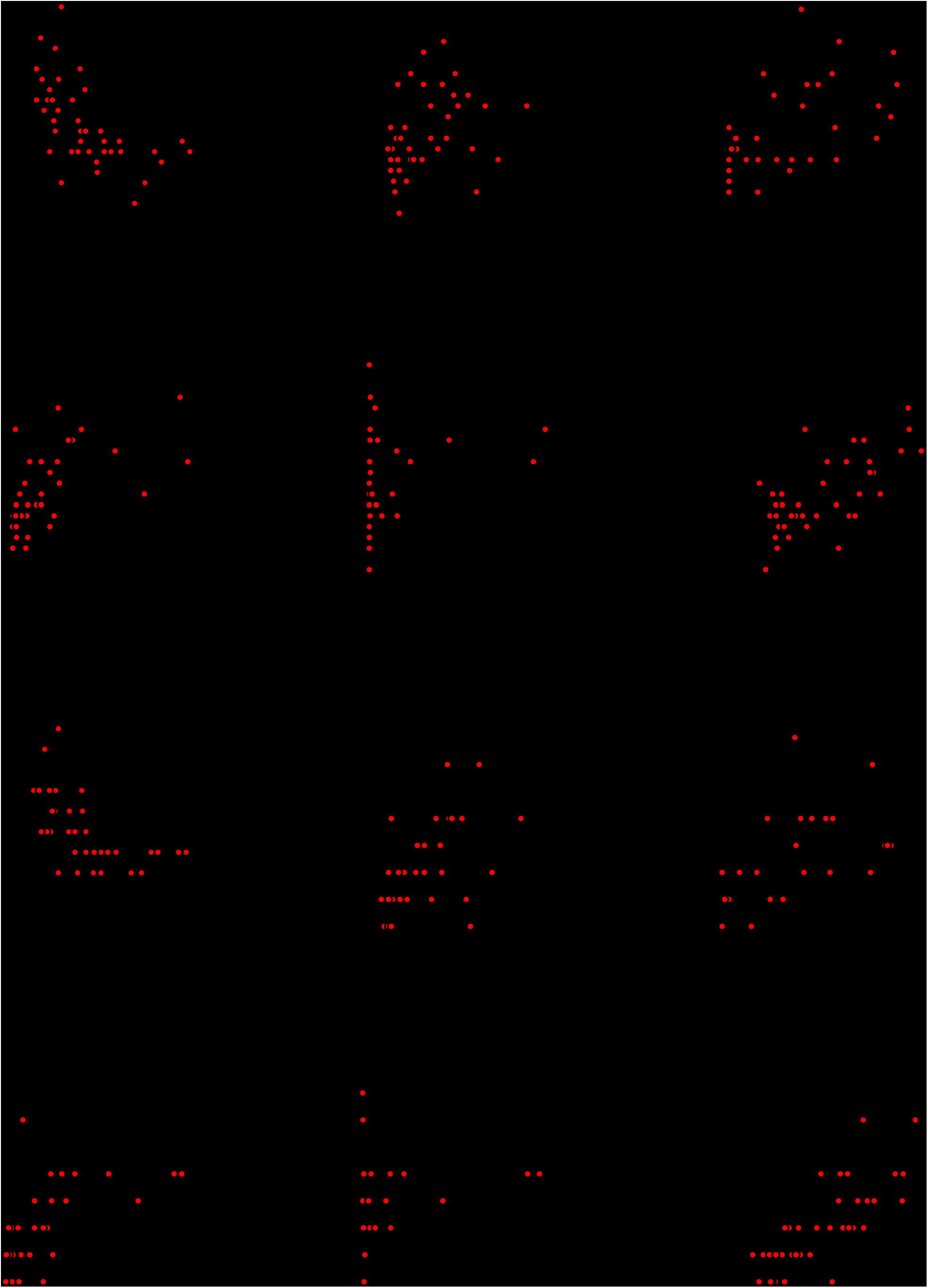

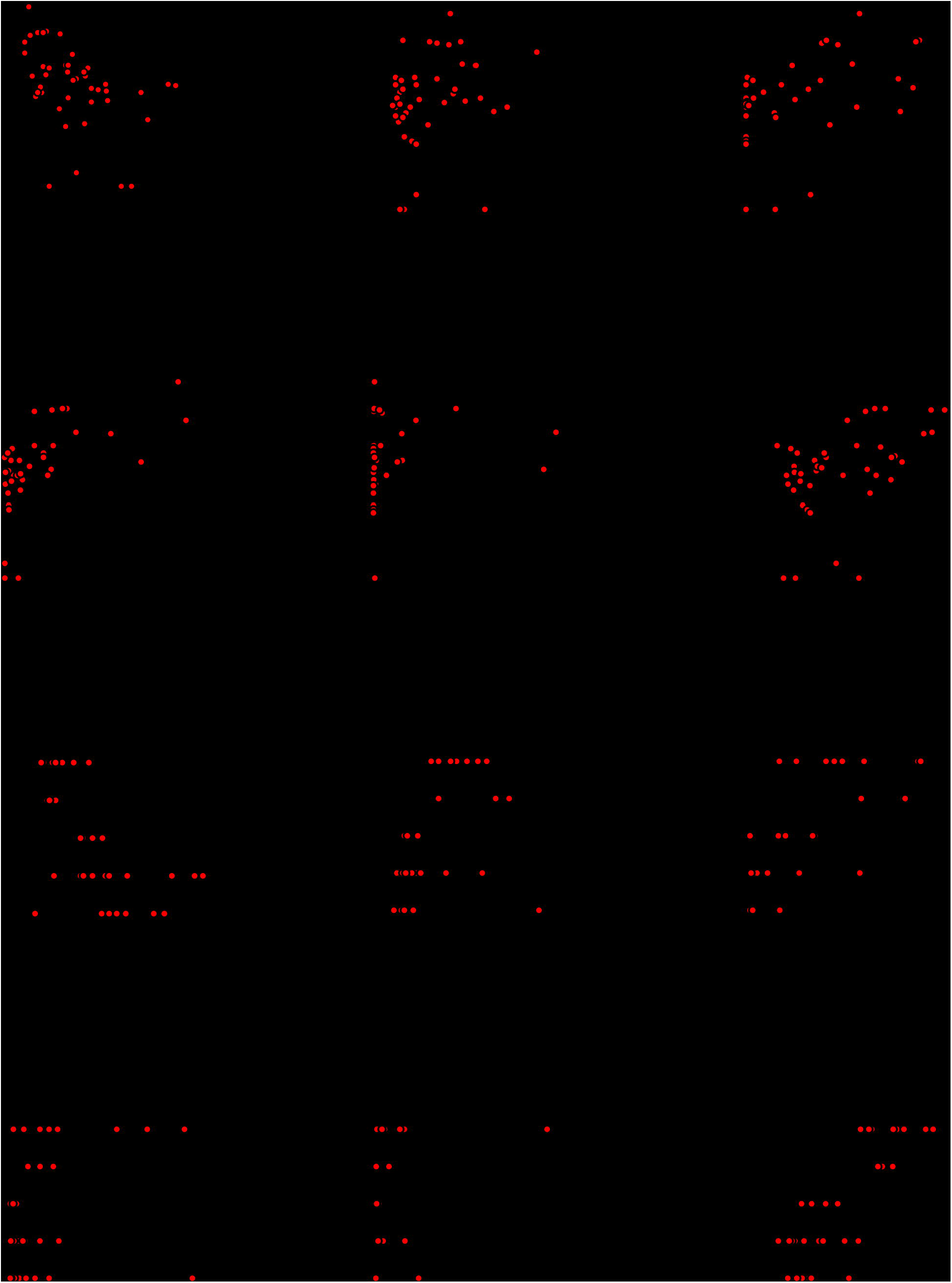

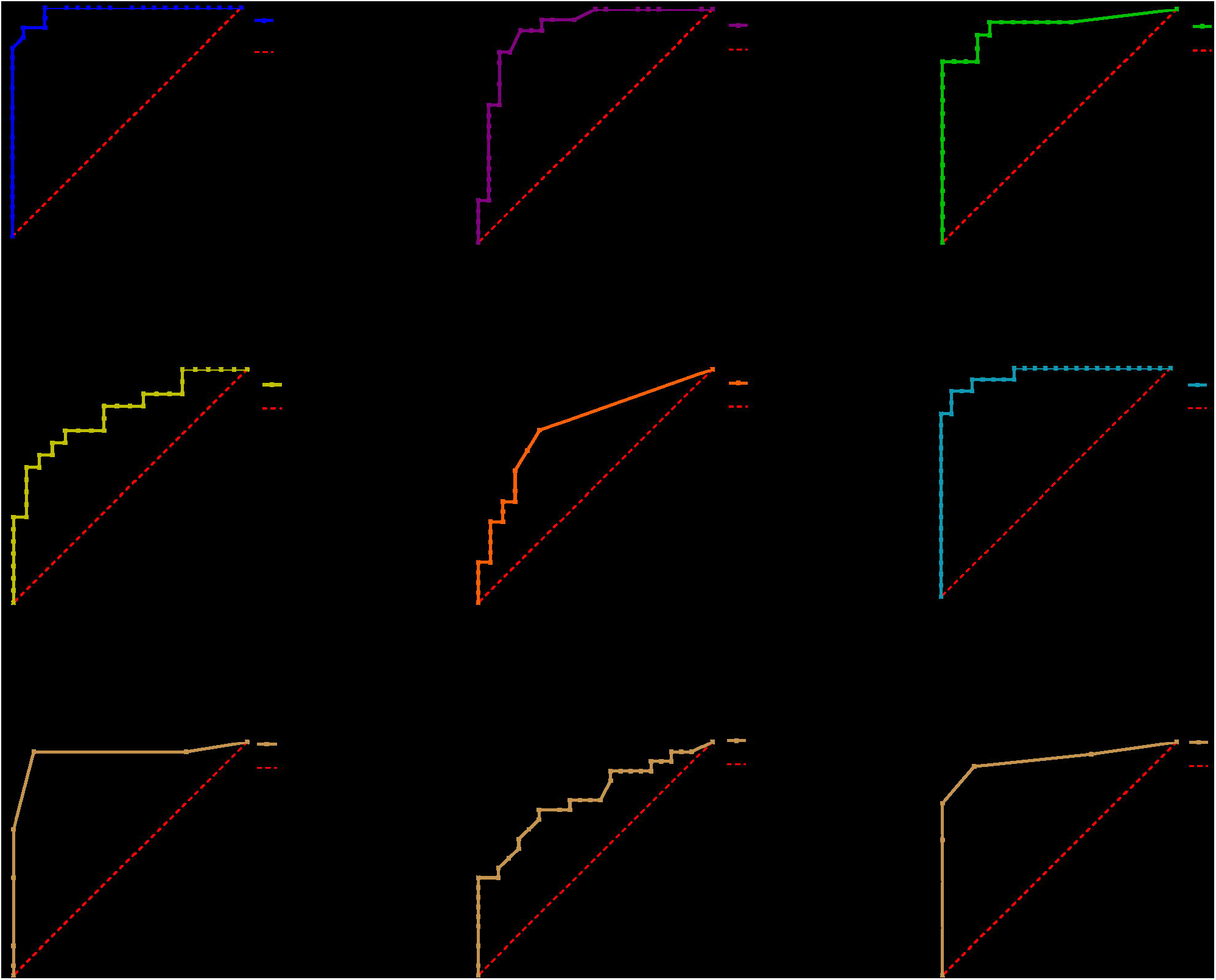

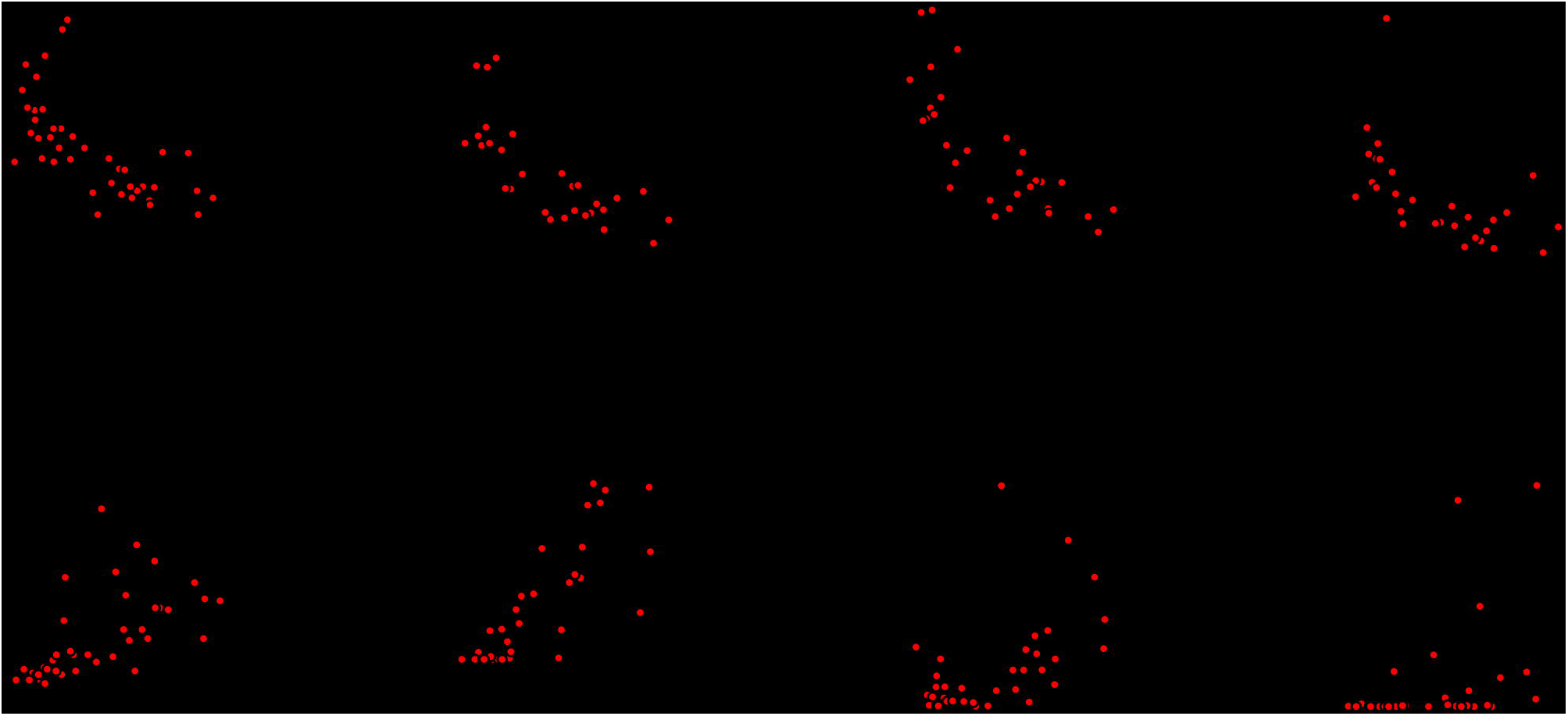

